# The Dichotomy of Progress: Public Perception, Scientific Evidence, and the Future of Global Health Governance—Insights from Georgia

**DOI:** 10.1101/2025.08.01.25332642

**Authors:** Zurab Alkhanishvili

## Abstract

**Background:** Despite significant advancements in global health science and governance, a troubling gap has emerged between objective progress and public perception. This disconnect—where life expectancy has doubled and child mortality has plummeted, yet public belief suggests regression—poses a critical threat to the legitimacy and efficacy of public health policy. This study measures the scope of this perception gap and examines its implications for the future of global health governance.

**Methodology/Principal Findings:** A nationally representative survey of 1,876 adults in Georgia (2023) revealed striking misconceptions: 64% believed global population growth had stagnated or reversed, 82% thought life expectancy had declined, and 88% incorrectly believed antibiotics are effective against viruses. In contrast, 98% of respondents accurately recognized the decline in child mortality. These findings demonstrate a substantial deficit in health literacy and reflect a broader misalignment between scientific progress and public understanding.

**Conclusions/Significance:** Bridging this disconnect demands a shift in how global health is communicated and governed. We propose a threefold strategy: investing in localized, trust-based public health communication; formally empowering youth as agents of change; and strategically leveraging digital media platforms to disseminate evidence-based knowledge. Scientific advancement without public trust is fragile—equitable and resilient global health governance in the 21st century depends on their alignment.

## 1. Introduction

The COVID-19 pandemic served as a stark reminder that infectious disease outbreaks are a recurring threat to humanity, profoundly testing the resilience and adaptability of global health systems. While unprecedented in its modern scale, this crisis follows a long history of epidemics, from the Spanish Flu to Ebola and SARS, that have consistently catalyzed significant shifts in global health governance. Past outbreaks have spurred crucial advancements, including the strengthening of the World Health Organization’s International Health Regulations after SARS, and the mobilization of massive international resources in response to the HIV/AIDS epidemic **[1].** Similarly, the COVID-19 pandemic led to remarkable feats of global cooperation, such as the accelerated development of vaccines, while simultaneously exposing deep-rooted vulnerabilities in our collective health infrastructure **[2]**.

However, this history of scientific and policy advancement reveals a troubling paradox: as progress in health security accelerates, a significant gap appears to be widening between empirical data and public perception. The very successes of public health—from eradicating diseases to extending life expectancy—have often become invisible to the societies they benefit. This disconnect became critically apparent during the COVID-19 pandemic, where scientific breakthroughs were often met with public skepticism and misinformation, thereby undermining the effectiveness of public health interventions **[3]**. This communication chasm represents a fundamental challenge to the future of global health governance.

To better understand and address this challenge, this paper poses a critical research question: to what extent does a gap exist between established scientific facts and public perception in a post-Soviet transitional society, and what are the broader implications for global health governance? To answer this, the primary aim of this study is to empirically investigate the scale of this disparity. To achieve this, we present original findings from a 2023 nationwide survey in Georgia, which assessed public knowledge on fundamental health metrics, including life expectancy, child mortality, and the efficacy of antibiotics. By analyzing these findings within the broader context of historical shifts in global health governance, this paper argues for a paradigm shift towards more localized, community-driven public health strategies. We highlight the underutilized potential of youth as agents of change and advocate for the strategic integration of modern digital platforms to bridge the critical gap between scientific progress and public trust, paving the way for a more resilient and participatory global health future.

## 2. Materials and Methods

To measure the scope of public misperceptions regarding foundational health indicators in Georgia, this study employed a quantitative survey methodology with a nationally representative sample.

### Survey Design and Sampling

The survey involved 1,876 respondents aged 18 and over from all 11 regions of Georgia, excluding the occupied territories, ensuring comprehensive geographic and demographic representation across both urban and rural settings. Data collection was conducted between March 1, 2023, and May 30, 2023. A probability sampling model, specifically cluster sampling with primary stratification, was adopted. The population was divided into 11 strata corresponding to the regions, with census areas serving as Primary Sampling Units (PSUs). Households within these units were selected as Secondary Sampling Units (SSUs) using the ‘random walk’ method, and the final respondent within the household was identified using the ‘last birthday’ method. This sample size afforded a margin of error of 2.5% at a 95% confidence level.

### Data Collection and Survey Focus

Data were gathered through face-to-face interviews using a structured, paper-based questionnaire that required approximately 3-5 minutes to complete. The questionnaire was designed to assess public knowledge and perceptions regarding four main indicators:

- Population growth: Whether respondents believed the global population has increased or decreased in the past century.
- Average life expectancy: Respondents’ views on whether life expectancy has risen or fallen over the last century.
- Child mortality rates: Public opinion on whether child mortality has increased or decreased.
- Efficacy of antibiotics: Whether respondents believed antibiotics are effective against viral infections such as the common cold, COVID-19, or influenza.

### Data Analysis

The collected responses were analyzed using SPSS statistical software. Descriptive statistics and cross-tabulations were employed to examine frequency distributions across key variables. Inferential statistical methods, including chi-square tests for categorical variables and t-tests for continuous variables, were used to assess the significance of observed differences. To ensure national representativeness and mitigate sampling bias, statistical weighting was applied. All analyses accounted for the survey’s complex sampling design, allowing for accurate population-level inferences.

### Ethics Statement

The study protocol was reviewed and approved by the Institutional Review Board (IRB) of the Health Research Union (IRB# 2023-00) on January 16, 2023. The Board determined that the study provided adequate safeguards for the rights and welfare of the participants. The study was conducted in accordance with the principles of the Declaration of Helsinki. All participants were provided with information about the study’s purpose and provided verbal informed consent to participate. The survey was conducted anonymously to ensure respondent confidentiality.

## 3. Results

The 2023 nationwide survey in Georgia revealed a profound disparity between public perceptions and established scientific data across several key health indicators. The findings highlight critical gaps in public knowledge concerning demographic trends and fundamental principles of medicine. A summary of the key findings is presented in Table 1.

**Table 1.** Discrepancies Between Public Perception and Factual Data on Key Health Indicators from a 2023 Nationwide Survey in Georgia (n=1,876)

| Health Indicator | Prevalent Public Misconception | Respondents Holding Misconception (%) | Factual Data |
| --- | --- | --- | --- |
| Global Population Growth | Global population has decreased or stagnated over the past century. | 64% | Increased from approx. 1.8 billion (early 20th C.) to over 8 billion. |
| **Average Life Expectancy** | Average life expectancy has decreased over the past century. | 82% | More than doubled globally since the early 20th century. |
| **Antibiotic Efficacy** | Antibiotics are effective against viral infections (e.g., common cold, COVID-19). | 88% | Ineffective against viruses; exclusively target bacterial infections. |
| **Child Mortality** | *Perception aligned with reality.* | *(2% held the misconception that it has increased)* | Dramatically decreased globally. |

### Public Perception of Demographic Trends

Regarding global population growth over the last century, a majority of respondents (64%) either believed the population has decreased or had no opinion. The primary reasons cited for the perceived decline were wars, pandemics, and environmental degradation. Only 36% of respondents correctly stated that the global population has increased.

A more pronounced gap was observed concerning life expectancy. An overwhelming 82% of respondents incorrectly believed that average life expectancy has decreased over the last century. The predominant reasons given included perceived deterioration of air and food quality and folk beliefs that their ancestors lived longer.

In contrast, public knowledge regarding child mortality was highly accurate. 98% of respondents correctly acknowledged that child mortality rates have decreased over the last century.

### Public Perception of Antibiotic Efficacy

The survey identified a critical misunderstanding of basic medical principles. When asked about the effectiveness of antibiotics for viral illnesses such as the common cold, COVID-19, or the flu, only 12% of respondents correctly stated that antibiotics are ineffective against viruses. Consequently, 88% of the surveyed population holds an incorrect belief regarding the fundamental mechanism and proper use of antibiotics.

Inferential analysis revealed no statistically significant differences in the prevalence of these misconceptions across key demographic variables such as age or urban/rural residence (p > 0.05), suggesting these knowledge gaps are widespread throughout the population

## 4. Discussion

The findings from our 2023 survey in Georgia reveal a profound disconnect between the objective successes of modern medicine and the subjective perceptions of the public. Georgia serves as a particularly compelling case study for examining these global trends. As a post-Soviet nation undergoing a protracted democratic transition, it reflects a common regional challenge of low public trust in state institutions, coupled with a healthcare system still grappling with the legacy of a centralized past **[4-5]**. This context creates a fertile ground for health-related misinformation and complicates the implementation of evidence-based public health communication. The chasm identified in our research is not merely a local curiosity but a powerful illustration of a critical vulnerability in contemporary global health governance. While the 98% accuracy in perceiving decreased child mortality suggests that some clear public health victories can penetrate public consciousness, the widespread misconceptions about population growth, life expectancy, and antibiotic efficacy underscore a systemic failure. The finding that 88% of Georgians misunderstand antibiotic efficacy is particularly alarming, yet it is not an isolated phenomenon. Similar trends have been documented globally, with significant percentages of individuals in various countries mistakenly believing that antibiotics can treat viral infections **[6-10].**

This paradox of progress, where achievements go unrecognized, is starkly evident when survey results are contrasted with the historical record. The global population has witnessed an extraordinary expansion, growing from approximately 1.8 billion in 1918 to over 8 billion today **[11]**. Concurrently, global average life expectancy has more than doubled, surging from around 32 years in the early 20th century to over 73 years by 2023 **[12]**. This monumental leap is the direct result of multifaceted advancements, including medical breakthroughs like antibiotics and vaccines **[13]**, agricultural innovations from the Green Revolution **[14]**, enhanced sanitation and transport infrastructure **[15]**, and broader access to education and nutrition **[16]**. The scale of this progress is best illustrated by comparing the scientific response to COVID-19 with that of the 1918 Spanish Flu. In 1918, the scientific toolkit was severely limited; in contrast, the rapid identification of the SARS-CoV-2 virus and the development of vaccines in record time showcased immense scientific advancement **[17]**. Yet, this progress is poorly understood, leading to the misuse of achievements like antibiotics and fueling the rise of antimicrobial resistance (AMR)—a premier global health security threat.

The communication gap is further exacerbated by the increasingly complex landscape of global health governance. The 20th-century model, primarily guided by UN agencies like the World Health Organization (WHO), UNICEF, and UNFPA, has transformed into a multifaceted arena. The HIV/AIDS crisis catalyzed the creation of critical entities like UNAIDS (1996) and The Global Fund to Fight AIDS, Tuberculosis, and Malaria (2002) **[18, 19]**. The 21st century saw the rise of influential European organizations like the ECDC (2005), powerful international coalitions like Gavi, the Vaccine Alliance (2000), and philanthropic giants such as the Bill and Melinda Gates Foundation **[20-21]**. The United States diversified its contributions through agencies like the CDC and targeted programs like PEPFAR (2003), while frontline NGOs such as Médecins Sans Frontières (MSF) took on essential roles **[22]**. While this diversity of actors brought immense resources, it also led to a fragmented system with conflicting responsibilities and disjointed communication efforts, making it exceedingly difficult to craft a single, clear public health message **[23-24]**.

Bridging the perception gap requires a fundamental shift in strategy, leading to several key policy recommendations. First, there is an urgent need to prioritize localized, community-centric health initiatives. Global strategies often fail without the active partnership of local NGOs and community leaders who can build trust and ensure interventions are culturally appropriate **[25]**. Second, policymakers must formally integrate youth as active partners in policy and implementation. With half the global population under 30, their role as digital natives and change agents is critical for future success. Third, a robust governance framework demands the strategic use of digital communication platforms. The failure to utilize platforms popular among youth during the COVID-19 pandemic represents a significant missed opportunity **[26-27]**. Finally, this all must be built on a foundation of transparency and inclusive, multi-stakeholder collaboration to foster a more resilient and responsive system **[28-29]**.

While this study’s focus on Georgia limits the generalizability of its specific quantitative findings, the discord it reveals between scientific data and public belief is a recognized global challenge, supported by evidence from various regions. Future research should expand this type of investigation across diverse cultural and economic settings to further map the contours of this problem. However, the path forward is clear: a resilient global health governance framework depends on its ability to effectively align scientific knowledge with public understanding and participation.

### Study Limitations and Future Directions

This study is based on a nationally representative survey conducted in Georgia, which may limit the generalizability of the findings to other countries with different sociopolitical, cultural, or health system contexts. While the patterns observed — particularly the gap between scientific progress and public perception — may resonate with broader global phenomena, they must be interpreted within the specific historical and institutional landscape of Georgia. Additionally, the study captures perceptions at a particular point in time; longitudinal research would be beneficial to assess how these attitudes evolve and respond to major global health events or policy changes.

Future research could expand on these findings by employing cross-national comparative designs, incorporating qualitative insights, and exploring the effectiveness of various communication strategies aimed at narrowing the perception gap. Integrating public opinion data with behavioral outcomes may also offer deeper understanding of the consequences of misalignment between perception and progress.

## 5. Conclusion

This study has illuminated the profound dichotomy that defines the current era of global health: while scientific capabilities and governance structures have made historic leaps forward, public understanding has not kept pace. The empirical findings from Georgia provide a stark, local snapshot of this global vulnerability, confirming that scientific progress without commensurate public trust is inherently fragile. The path forward, therefore, is not only through further innovation but through a fundamental rethinking of public health engagement. Ultimately, global health governance can only meet the challenges of the 21st century if scientific progress is matched by public trust and participation.

## Data Availability

All relevant data from this study are included within the article. The survey was based on a simple questionnaire, and all resulting aggregate data (frequencies and percentages) are presented in the Results section and Table 1 of the manuscript. The minimal, anonymized dataset is available from the corresponding author upon reasonable request. It should be noted that the original data were collected in the Georgian language.

